# Early but not late convalescent plasma is associated with better survival in moderate-to-severe COVID-19

**DOI:** 10.1101/2021.06.16.21258972

**Authors:** Neima Briggs, Michael V. Gormally, Fangyong Li, Sabrina L. Browning, Miriam M. Treggiari, Alyssa Morrison, Maudry Laurent-Rolle, Yanhong Deng, Jeanne E. Hendrickson, Christopher A. Tormey, Mahalia S. Desruisseaux

## Abstract

**Background:** Limited therapeutic options exist for coronavirus disease 2019 (COVID-19). COVID-19 convalescent plasma (CCP) is a potential therapeutic, but there is limited data for patients with moderate-to-severe disease.

**Research Question:** What are outcomes associated with administration of CCP in patients with moderate-to-severe COVID-19 infection?

**Study Design and Methods:** We conducted a propensity score-matched analysis of patients with moderate-to-severe COVID-19. The primary endpoints were in-hospital mortality. Secondary endpoints were number of days alive and ventilator-free at 30 days; length of hospital stay; and change in WHO scores from CCP administration (or index date) to discharge. Of 151 patients who received CCP, 132 had complete follow-up data. Patients were transfused after a median of 6 hospital days; thus, we investigated the effect of convalescent plasma before and after this timepoint with 77 early (within 6 days) and 55 late (after 6 days) recipients. Among 3,217 inpatients who did not receive CCP, 2,551 were available for matching.

**Results:** Early CCP recipients, of whom 31 (40%) were on mechanical ventilation, had lower 14-day (15% vs 23%) and 30-day (38% vs 49%) mortality compared to a matched unexposed cohort, with nearly 50% lower likelihood of in-hospital mortality (HR 0.52, [95% CI 0.28-0.96]; *P*=0.036). Early plasma recipients had more days alive and ventilator-free at 30 days (+3.3 days, [95% CI 0.2 to 6.3 days]; *P*=0.04) and improved WHO scores at 7 days (−0.8, [95% CI: −1.2 to - 0.4]; *P*=0.0003) and hospital discharge (−0.9, [95% CI: −1.5 to −0.3]; *P*=0.004) compared to the matched unexposed cohort. No clinical differences were observed in late plasma recipients.

**Interpretation:** Early administration of CCP improves outcomes in patients with moderate-to-severe COVID-19, while improvement was not observed with late CCP administration. The importance of timing of administration should be addressed in specifically designed trials.

## Introduction

Severe acute respiratory syndrome coronavirus 2 (SARS-CoV-2), a positive-sense, single-stranded RNA virus of the *Coronaviridae* family, is the etiologic agent of Coronavirus disease 2019 (COVID-19) and is responsible for greater than 126 million global infections and approximately 2.7 million deaths since December 2019.[1-3] During the first 6 months of the COVID-19 pandemic, treatment was largely supportive. In June 2020 the Randomized Evaluation of Covid-19 Therapy (RECOVERY) trial reported that dexamethasone reduced mortality in patients on respiratory support; thus, corticosteroids have become standard of care for patients on supplementary oxygen.[4, 5] Furthermore, remdesivir, a nucleotide analogue that disrupts viral replication, is the only other pharmacologic approved for COVID-19 in the United States [6], although with little to no effect on mortality or clinical course in hospitalized patients.[7] No additional therapeutics have yet to show efficacy in hospitalized patients with COVID-19.

Convalescent plasma containing antibodies generated following pathogens’ exposure has been employed in past epidemics as a means of passively transferring immunity from individuals with resolved infection. In addition to antibody-mediated protection via neutralization, therapeutic antibodies can directly induce cellular cytotoxicity, complement activation, and phagocytosis. Furthermore, convalescent plasma may contain beneficial anti-inflammatory cytokines, defensins, and pentraxins that quell a severe inflammatory response.[8] During the SARS [9] epidemic in 2003 and more recently during the H1N1[10] pandemic of 2009, treatment with convalescent plasma resulted in significant mortality reduction.

In the current pandemic, single-arm observational studies have reported administration of COVID-19 convalescent plasma (CCP) to patients with mild to severe COVID-19 with variable results and limited cohort sizes.[11-15] Two randomized open-label trials were terminated prior to full enrollment; one due to drop in local COVID-19 incidence and a second due to concerns that the recruited cohort had preexisting anti-SARS-CoV-2 antibodies prior to enrollment.[16, 17] A third, randomized open-label trial of 464 hospitalized adults with moderate COVID-19 failed to show benefit in 28-day mortality or progression to severe disease; however, the study was limited by low or undetectable antibody titers in the donor plasma [18]. Most recently, a randomized, double-blinded, placebo-controlled trial found that administration of higher-titer CCP within 72 hours after symptom onset among mildly infected older adults reduced risk of progression to severe respiratory disease by 48%.[19]

There is increasing evidence that clinical response to CCP might depend on the timing of administration. We conducted a propensity score matched analysis in one of the largest reported cohort of hospitalized patients with moderate-to-severe COVID-19 to investigate outcomes associated with early versus late transfusion of CCP.

## Methods

### Study Design

This was a cohort study that included 3,368 patients hospitalized with moderate (supplemental oxygen flow rate of ≥3L/min) to severe or life-threatening (respiratory failure, shock, or multi-organ failure) COVID-19 managed within the Yale New Haven Health system (YNHHS) from March 8, 2020.to July 25, 2020. Of those, a total of 151 patients were transfused with CCP. We dichotomized the cohort based on the median time to CCP administration (6 days; interquartile range: 3, 11 days): The early CCP cohort received treatment within 6 days of hospitalization, while the late CCP cohort received treatment after 6 days of hospitalization. This cut-off was consistent with transfusion time used in another study.[15] For the early or late CCP cohorts, we conducted distinct propensity score matchings to generate two 1:1 matched unexposed cohorts, as previously described.[20] Index dates for the unexposed patients were assigned corresponding to the day of CCP administration within each matched pair.

This study was approved by the Yale University Institutional Review Board (HIC#: 2000027871) with a waiver of informed consent.

### Eligibility and Selection of Convalescent Plasma Recipients

In April 2020 the U.S. Food and Drug Administration (FDA) provided physicians access to convalescent plasma through the US National Expanded Access Program (EAP), which was led by the Mayo Clinic (IRB# 20-003312). Patients hospitalized with COVID-19 within YNHHS were screened for eligibility to receive CCP through the EAP based on shared decision making between the Convalescent Plasma Clinical Team, primary medical team, and the patient or Legally Authorized Representative. Patients were eligible to receive CCP if they were 18 years of age or older, had a laboratory confirmed diagnosis of SARS-CoV-2 infection, were hospitalized within YNHHS with COVID-19 complications, and had moderate-to-severe disease as indicated by the following characteristics: 1. Supplementary oxygen requirements (≥3L/min nasal cannula) with pulmonary infiltrates on chest imaging; 2. Refractory acute respiratory failure; 3. Septic shock; or 4. Multisystem organ dysfunction. Absolute contraindication to the administration of CCP was confirmed new thromboembolic event. Relative contraindications were: 1. Confirmed or high suspicion for bacterial or fungal infection; 2. Suspicion of a new thromboembolic event; 3. Recent significant hemorrhage; 4. High risk for hemorrhage and on therapeutic anticoagulation; and 5. Severe IgA deficiency. At least two physicians on the Convalescent Plasma Clinical Team reviewed each CCP request and approved them based on the adherence to the inclusion criteria and absolute or relative contraindications.

### Convalescent Plasma Donors

CCP was obtained from the hospital system’s blood suppliers, including New York Blood Center (New York, NY), Rhode Island Blood Center (Providence, RI), and the American Red Cross (Farmington, CT). Over the course of the study period the criteria for eligibility for CCP donation was modified by the FDA, though generally required PCR-confirmed diagnosis of COVID-19 and complete resolution of symptoms for greater than 14 days prior to donation. The units transfused were not labeled by the blood suppliers with an anti-SARS-CoV-2 IgG titer.

### Preparation and Administration of Convalescent Plasma

Each patient received one unit of ABO compatible CCP, with a typical unit being approximately 200 mL in volume. Units were thawed on demand once requested and the patient was confirmed to have met all eligibility criteria.

### Outcome measures

The primary endpoint was in-hospital mortality. Time was censored on the date of discharge or administratively on August 3, 2020, the date of closure of data extraction. Secondary endpoints included days alive and free from mechanical ventilation 30 days post-index date, length of hospital stay post CCP or index date in the unexposed cohort, and change in 8-point WHO ordinal scale at 7 days post CCP or index date and at hospital discharge (**S1 Table**).

### Statistical Analysis

Patient demographics and pre-existing comorbidities were collected at admission. Vital signs, oxygen therapy, concomitant medications, and laboratory tests were collected longitudinally during the hospitalization. Descriptive statistics were used to summarize patient demographics and clinical characteristics. In bivariate analysis, two-sample Student’s t-test, Wilcoxon rank-sum test, Chi-square or Fisher’s exact tests were used for comparisons of exposed and unexposed cohorts, as appropriate.

Two separate propensity-score matches were carried out to select unexposed patients for the early CCP cohort or unexposed patients for the late CCP cohort (**S2 Figure and S3 Figure**). CCP recipients were matched with unexposed patients using both propensity scores and exact matching on the worst WHO scores preceding CCP administration or index date, and on administration of remdesivir. No replacement was allowed for matching, and matched unexposed patients in the early cohort were allowed in the late cohort. The standardized mean difference between matched groups was set at ≤ 0.25. Because of the stringent exact matching criteria, the optimal algorithm that minimized differences between the matching pairs was only able to yield 1:1 unexposed match. Group comparisons were performed to confirm expected balance between matched groups. For each matched cohort, a multivariate logistic regression model was fitted to estimate the probability of receiving CCP within the specified time frame and create propensity for each individual (**S2 Table**). The models included demographic and clinical characteristics at hospital admission; concomitant medications, laboratory results prior to CCP administration, and supplemental oxygen status as covariates. The list of covariates was presented in S2 Table.

For the primary analysis, we used the Kaplan-Meier product-limit estimator to estimate the hospital mortality function. We compared CCP exposed and unexposed patients for the early and for the late cohorts using the log-rank test. Mortality at 14 days and 30 days post CCP or index date was estimated and reported with 95% confidence intervals. Secondary outcomes were analyzed using mixed effects models or ordinal logistic regression with robust variance estimation to compare treatment groups with their respective matched unexposed cohort. The effect sizes are presented as mean difference with 95% confidence interval. A two-sided alpha level of 0.05 was required for statistical significance. All statistical analyses were performed using SAS 9.4 (Cary, NC) software.

## Results

Of 3,368 patients with confirmed SARS-CoV-2 infection within YNHHS, 151 eligible patients received CCP under the US National Expanded Access Program (**Figure 1**). Among the CCP recipients, 80 received CCP within 6 days after admission, and 71 received CCP after 6 days of hospitalization. Of those who received CCP, 3 early and 16 late recipients were excluded from propensity score matching due to missing laboratory values and/or supplemental oxygen information during the first week of hospitalization, leaving a total of 77 CCP patients in the early cohort and 55 CCP patients in the late cohort available for inclusion in the analyses. One of three excluded early recipients died (33%), and 9 out of 16 excluded late recipients died (56.3%). Among 3,217 SARS-CoV-2 positive hospitalized patients who did not receive CCP transfusion, 666 were excluded from the unexposed cohort due to incomplete clinical data, leaving 2,551 patients eligible for propensity score matching.

**Figure 1:**
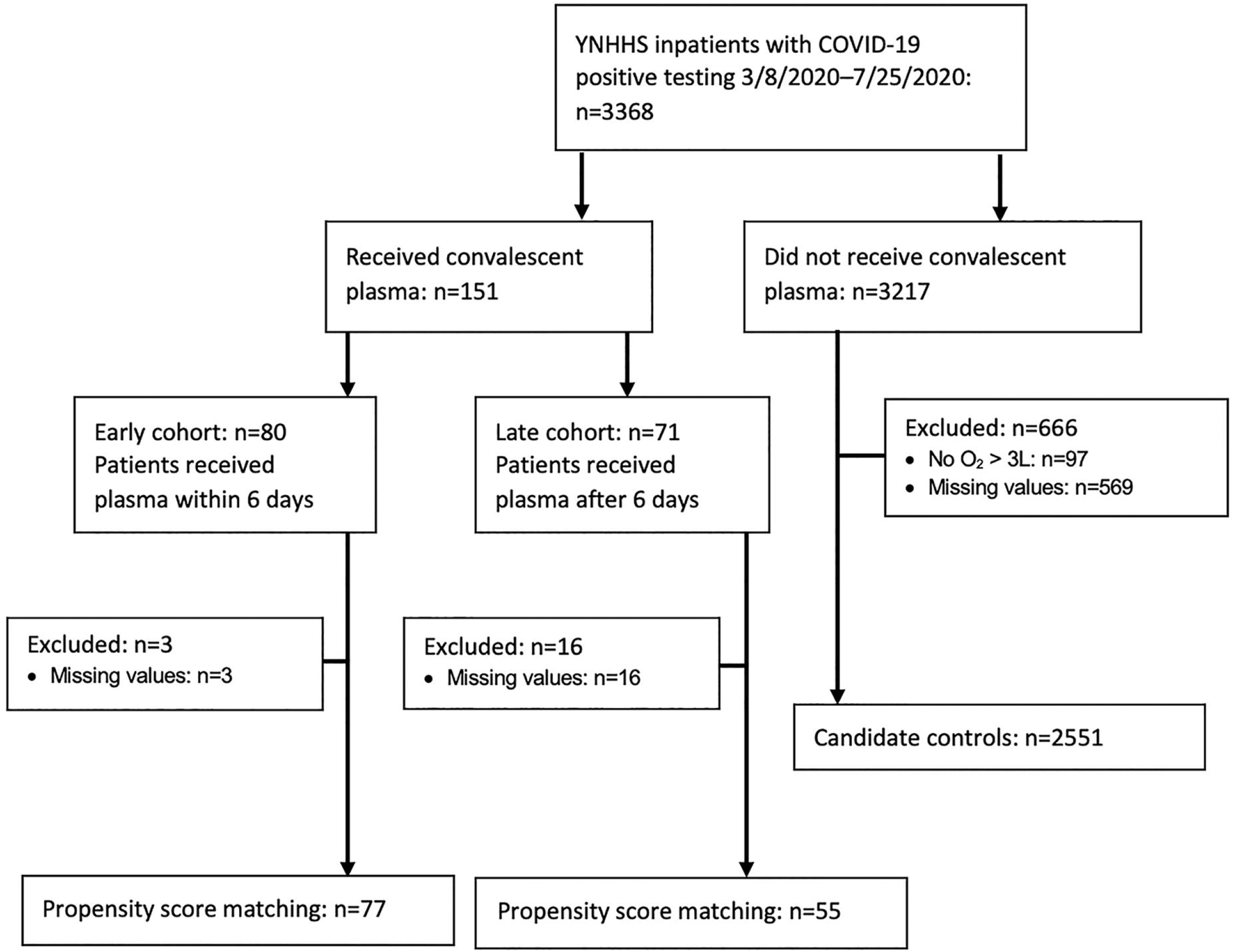
Consort selection tree. Patient disposition in the observational cohort analysis.

Baseline demographic characteristics and clinical factors for the early and late cohorts are summarized in **Table 1**. Overall, age, sex, race, ethnicity, BMI, and Charlson Comorbidity Index were equally distributed between the CCP and the unexposed group. Likewise, the mean maximum WHO score prior to the index date and the level of supplemental oxygen were comparable between CCP and unexposed group. Baseline laboratory values obtained prior to CCP administration or index date were similar among the CCP and unexposed group, with the exception of mean fibrinogen level which was higher in the CCP treated group compared with the unexposed group in the late cohort. Concomitant medications, including hydroxychloroquine, remdesivir, tocilizumab, and dexamethasone were well balanced in the early cohort. Tocilizumab was administered more frequently in the CCP group compared with the unexposed group in the late cohort (**Table 1**).

**Table 1:**
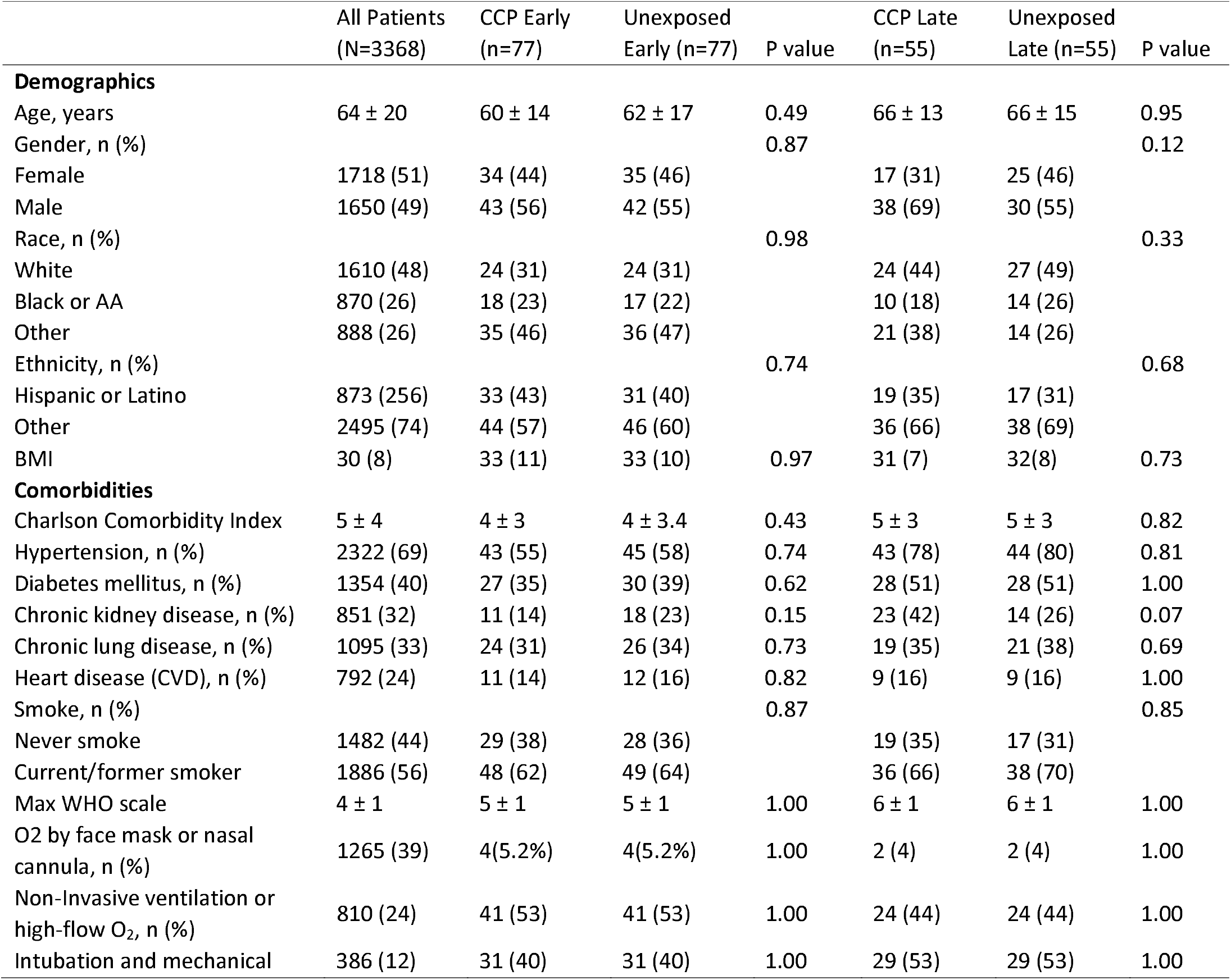
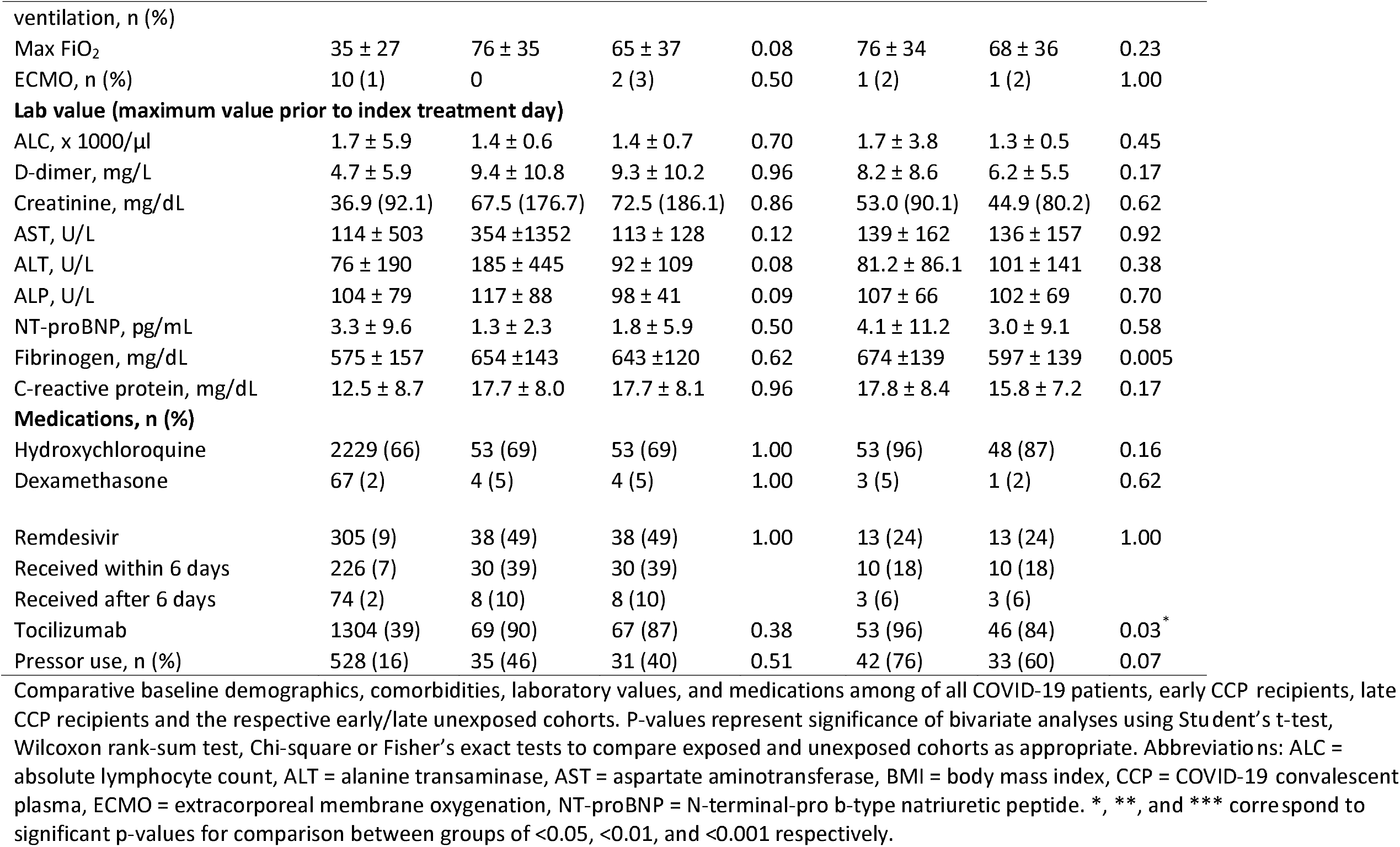
Baseline demographic characteristics and clinical factors.

### Primary Endpoint

In the propensity score matched analysis, the estimated in-hospital mortality was 15% by day 14 among early CCP recipients, compared with 23% in their matched unexposed cohort. At day 30, estimated in-hospital mortality was 38% in early CCP recipients compared to 49% in their matched unexposed cohort (**Table 2**). Overall, early CCP recipients were nearly 50% less likely to die in the hospital compared to patients in the matched unexposed cohort (HR 0.52, [95% CI 0.28 to 0.96]; *P*=0.036) (**Figure 2a**). In contrast, there were no differences in mortality among late CCP recipients compared to their matched unexposed cohort (HR 0.98, [95% CI 0.53 to 1.83]; *P*=0.95) (**Figure 2b, Table 2**). In the late cohort, the estimates of 14-day and 30-day in-hospital mortality were 28% vs 29% and 42% vs 47%, comparing CCP recipients vs matched unexposed patients, respectively.

**Table 2:** Primary and secondary study endpoints.

|  | CCP Early | Unexposed Early | Difference or<br>HR (95% CI) | p-value | CCP Late | Unexposed Late | Difference or<br>HR (95% CI) | p-value |
| --- | --- | --- | --- | --- | --- | --- | --- | --- |
| **Primary outcomes** |  |  |  |  |  |  |  |  |
| 14-day mortality | 15% | 23% |  |  | 28% | 29% |  |  |
| 30-day mortality | 38% | 49% | 0.52 (0.28,<br>0.96) | 0.036* | 42% | 47% | 0.98 (0.53, 1.83) | 0.95 |
| **Secondary outcomes** |  |  |  |  |  |  |  |  |
| Alive and mechanical<br>ventilation-free, days (SD) | 21.4 (11.6) | 18.1 (12.6) | 3.3 (0.2, 6.3) | 0.04* | 13.4 (12.3) | 16.9 (12.8) | -3.5 (-8.6, 1.7) | 0.18 |
| Length of hospitalization,<br>days (95% CI) | 18.4 (16.3, 20.7) | 17.5 (14.9, 20.1) | 1.0 (-2.4,4.4) | 0.57 | 24.7 (22.6, 26.8) | 19.9 (16.9, 22.9) | 4.9(1.2, 8.5) | 0.01* |
| WHO score on index<br>date, mean (SD) | 5.2 (0.7) | 4.9 (0.9) | 0.3 (0.2,0.5) | 0.0002*** | 5.7 (0.5) | 5.3 (0.9) | 0.4 (0.1, 0.6) | 0.0001*** |
| WHO score change at 7-days, mean (SD) | -0.5 (1.1) | 0.3 (1.6) | -0.8 (-1.2, -0.4) | 0.0003*** | 0 (1.3) | -0.2 (1.6) | 0.2 (-0.3, 0.8) | 0.41 |
| Change of WHO score at<br>discharge, mean (SD) | -0.7 (2.0) | 0.2 (2.2) | -0.9 (-1.5, -0.3) | 0.004** | -0.1 (2.3) | -0.4 (2.1) | 0.6 (-0.4, 1.5) | 0.23 |

Table reports primary and secondary outcomes for the two separate propensity score matched cohorts of early and late transfused patients. CCP effect on mortality is reported as a hazard ratio and secondary outcomes report differences in days or WHO score, each with 95% confidence intervals and p-values shown. Differences in alive and mechanical ventilation free days are measured at 30-days post index. Length of hospitalization is measured from index day. \*, \*\*, and \*\*\* correspond to significant p-values for comparison between groups of <0.05, <0.01, and <0.001 respectively.

**Figure 2:**
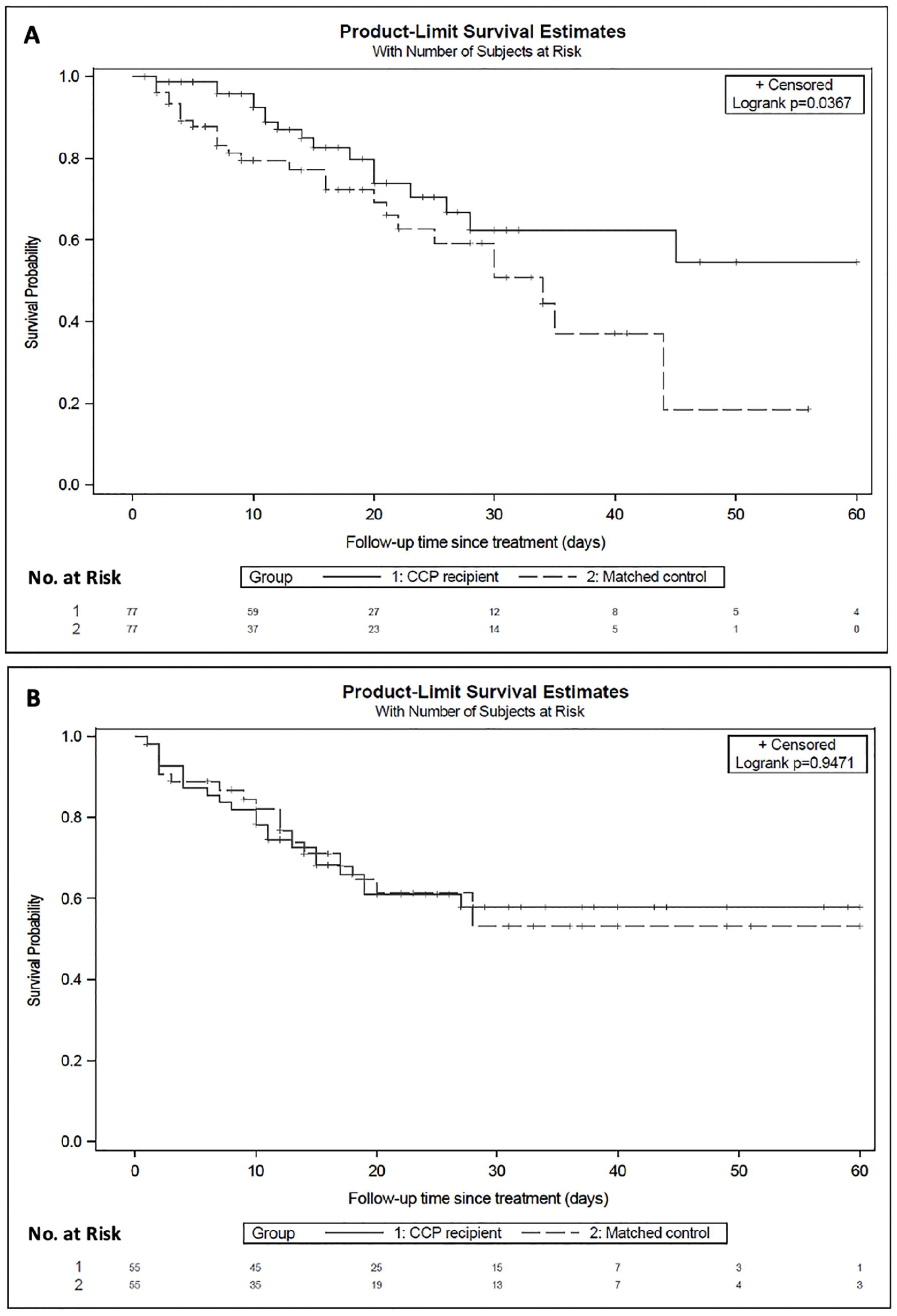
Kaplan-Meier curves of survival in early (A) and late (B) convalescent plasma recipients vs respective matched unexposed patients. Shown are Kaplan-Meier estimates of survival from time of index in (A) early CCP recipients (solid line) vs matched unexposed patients (dashed line) (B) late CCP recipients (solid line) vs matched unexposed patients (dashed line). Survival improved for early CCP at 14-days (15% vs 23%) and 30-days (38% vs 49%) compared to matched unexposed patients (HR 0.52, [95% CI 0.28-0.96]; p=0.0367). There was no difference in mortality at 14-days (28% vs 29%) or 30-days (42% vs 47%) among late CCP recipients compared to their matched unexposed patients (HR 0.98,[95% CI 0.53to 1.83]; p=0.95). Censoring is indicated by the tick mark “+” with number by each ten-day interval marked below the number at risk.

### Secondary Endpoints

The early CCP cohort had more days “alive and ventilator free” by 30 days post-index date compared with their matched unexposed cohort (mean 3.3 days, [95% CI 0.2 to 6.3]; *P*=0.04, **Table 2**). There were no differences comparing CCP recipients and matched unexposed patients in the late cohort (mean −3.5 days, [95% CI −8.6 to 1.7]; *P*=0.18).

In the early CCP cohort, WHO scores were significantly lower at 7 days post administration or post index date (difference in means −0.8, [95% CI −1.2 to −0.4]; *P*=0.0003) and at hospital discharge (difference in means −0.9, [95% CI −1.5 to −0.3]; *P*=0.004) compared to their matched unexposed cohort. Early CCP recipients were nearly twice as likely to demonstrate an improvement in their WHO scores at discharge from baseline index day scores (OR 1.9, [95% CI 1.1 to 3.3]; *P*=0.02, **Figure 3a, Table 2**). 42.9% of CCP recipients on mechanical ventilation at baseline (WHO score of 6) no longer required supplementary oxygen at the time of discharge (WHO score of 3) compared to 36.4% of unexposed matches (**S1 Figure**). There were no differences in length of hospital stay post treatment or index date (mean, 18.4 days vs 17.5 days, *P*=0.57) or likelihood of discharge (HR for discharge 0.9, 95% CI 0.6 to 1.3; *P*=0.46).

**Figure 3:**
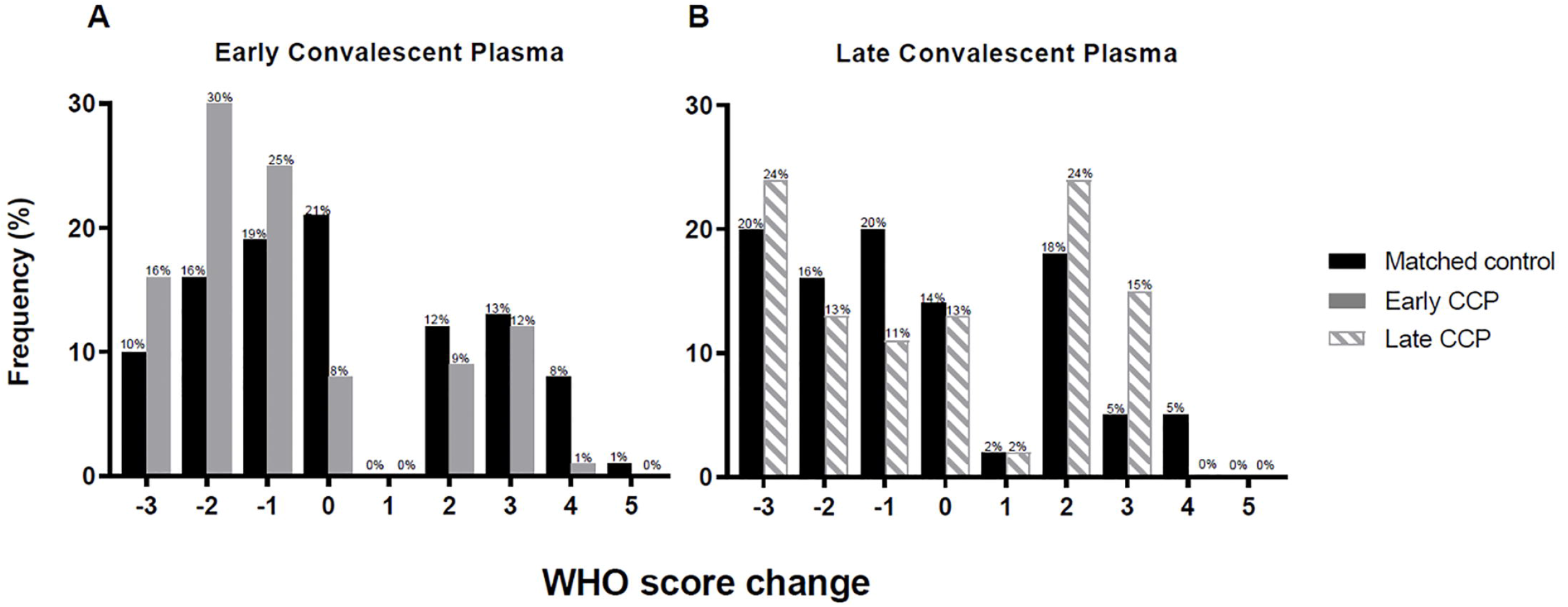
Histogram plots of WHO ordinal scale scores before and after intervention in the early (A) and late CCP (B) compared to matched unexposed cohorts. Shown is a histogram of the change is WHO COVID-19 severity score from time of index to discharge in (A) early CCP recipients (gray solid bars) vs matched unexposed patients (black solid bars) (B) late CCP recipients (gray dashed bars) vs matched unexposed patients (black solid bars). WHO scores in (A) early CCP recipients were significantly improved at discharge (difference in mean −0.9, [95% CI −1.5 to −0.3]; p=0.004) when compared to unexposed patients. Relative to the unexposed patients, early CCP recipients were nearly twice as likely to demonstrate an improvement in their WHO scores at discharge from baseline index day scores (OR 1.9,[95% CI 1.1 to 3.3]; p=0.02). Among (B) late CCP recipients, there was no effect of CCP on WHO scores at discharge (difference in mean +0.6, [95% CI −0.4 to 1.5]; p=0.23). The odds ratio for improvement at discharge was 0.6 (95% CI 0.3 to 1.3; p=0.22).

In the late cohort, there were no differences in WHO scores at day 7 (mean +0.2, [95% CI −0.3 to 0.8]; *P*=0.41) or at discharge (mean +0.6, [95% CI −0.4 to 1.5]; *P*=0.23) between CCP recipients and their matched unexposed cohort (**Figure 3b, Table 2**). The change from baseline (CCP transfusion or index date) to discharge in WHO scores was not different either (OR 0.6 [95% CI 0.3 to 1.3]; *P*=0.22). Hospital length of stay was longer in CCP recipients compared with unexposed matches, (mean, 24.7 days vs 19.9 days, *P*=0.01), although the likelihood of discharge was not different (HR for discharge 0.6, 95% CI 0.3 to 1.1) (**Table 2**).

### Adverse Outcomes

No immediate transfusion-related adverse events were observed after CCP administration in any of the cohorts and no transfusion reactions were reported to the blood bank.

## Discussion

In this cohort study of CCP transfusion in patients with moderate-to-severe COVID-19, we found a nearly 50% lower in-hospital mortality, fewer days spent on mechanical ventilation, and greater improvement in WHO scores after early administration of CCP (within 6 days of hospitalization) when compared to a propensity score-matched unexposed cohort. When administered after 6 days of hospitalization, CCP transfusion was not associated with improved mortality or clinical outcomes.

On August 23, 2020, the FDA granted Emergency Use Authorization for the use of CCP in the management of COVID-19 based on preliminary analysis of the EAP which found that CCP administered within 3 days of admission produced clinical benefit. Specifically, the observational study reported by Joyner et al. found lower 30-day mortality in patients who received plasma within 72 hours of first positive SARS-CoV-2 PCR compared to patients transfused after 72 hours.[14] There is a growing body of literature supporting a biological rationale for the early administration of CCP as a means to enhance response to this treatment.[15, 21-24] Active viral shedding is highest within the first week of symptoms, with peak replication occurring within the first 5 days.[21, 22] Moreover, antibody responses to SARS-CoV-2 start to develop in the first week, with antiviral immunoglobulin peaks by day 22 of symptoms.[23] With the SARS epidemic of 2003, Cheng et. al. described higher hospital discharge and lower mortality in patients who received anti-SARS convalescent plasma prior to 14 days after the onset of illness.[9] This is also consistent with data observed by Libster and colleagues, who found a 48% relative risk reduction in developing severe respiratory disease in older adults receiving CCP within 72 hours after the onset of mild COVID-19 symptoms.[19] However, this aforementioned study failed to detect any benefit in mortality due to very few deaths. Another small study using propensity score matched method reported benefit for patients with severe COVID receiving CCP within 7 days with a HR of 0.34.[15] The results of our investigation support these findings utilizing a larger sample size and suggest that the administration of CCP after the first week of hospitalization does not attain the same results as earlier CCP administration.

In contrast, there have been multiple randomized controlled trials of CCP with negative findings, the most recent being the CCP arm of RECOVERY, which found no benefit in mortality, mechanical ventilator liberation or hospital discharge.[25] An important distinction from the present study is that the median time to CCP in RECOVERY was 9 days from symptoms. Zeng and colleagues reported that administration of CCP 21 days after initial SARS-CoV-2 detection failed to improve survival despite successful viral clearance.[26] Simonovich et al. also reported no benefit to CCP in patients who had a median time from symptoms onset to CCP transfusion of 8 days in a recent study.[27] Indeed, our findings in the late cohort are consistent with this prior work and further emphasize the importance of early administration of CCP to achieve clinical improvement.

It is important to note that 40% of early CCP recipients in our cohort were on mechanical ventilation, suggesting that the clinical benefits of early CCP administration extend to even patients who are critically ill. Although previous uncontrolled case series reported improvement in mechanically ventilated patients who received CCP, [28, 29] an open-label, randomized trial by Li et. al. failed to demonstrate any significant clinical improvement in a small subset of patients with life-threatening COVID-19,[16] although fewer patients in the CCP group died compared to unexposed patients.[16] To our knowledge, our study is among the first to report significant clinical benefit with early administration of CCP in hospitalized patients with moderate-to-severe COVID-19.

Interestingly, in our study late CCP recipients had longer lengths of hospitalization than their matched unexposed counterparts despite having similar likelihood of discharge, mortality, and WHO scores at day 7 and discharge. Future studies should help elucidate potential explanations for these observations.

### Limitations

The interpretation of our findings is limited by the observational nature of the study. However, propensity score matching to parse electronic medical record data is a valid and commonplace method in medical research.[20, 30] Matching characteristics were comparable between exposed and unexposed patients. We acknowledge that it is possible that despite the adequate matching, patients in the late cohort were intrinsically different from matched unexposed patients, and indication bias might have played a stronger role, or the late administration of CCP might not be an adequate rescue intervention. While results from multi-center, randomized, controlled trials will be available in the future, our study is the first adequately powered to evaluate CCP for use in moderate-to-severe disease.

Although other studies may use different cut-offs for timing of CCP administration, our study used hospitalization date as a reliable time point that could be easily implemented to guide hospital treatment algorithms. It would be more challenging to establish timing of treatment based on the date of symptom onset, because initial COVID-related symptoms are often more difficult to precisely identify, and even more so in the early stages of the pandemic. Symptom onset information was also not feasible to obtain for our entire 2,551 patient cohort used for propensity matching. Another limitation of the present study is that each pool of plasma was unique without individual titer levels available, given that data for this study were collected prior to a validated serologic test. This information on donor plasma would have enabled us to confirm a dose-response relationship, supporting the concept of the biological effects of CCP.

## Conclusion

Among patients hospitalized with moderate-to-severe COVID-19, administration of CCP within 6 days of hospitalization in conjunction with standard of care treatment was associated with improved in-hospital mortality compared to standard of care treatment alone. Furthermore, the early administration of CCP was associated with increased 30-day alive and ventilator free days and improved WHO ordinal scale scores compared with the matched unexposed cohort. However, these associations were not observed among patients transfused 7 days or later into their hospital course. Further clinical trials are required to confirm the efficacy and safety of CCP, especially considering the importance of timing of administration, in order to address the limitations of this propensity-matched analysis and confirm our findings.

## Data Availability

All data contained herein are available accordingly.

## Abbreviations

ALC: Absolute lymphocyte count
AA: African American
ALT: Alanine transaminase
AST: aspartate aminotransferase
BMI: Body mass index
CCI: Charlson cormorbidity index COVID-19 Coronavirus disease 2019
CCP: COVID-19 convalescent plasma
EAP: Expanded Access Program
ECMO: Extracorporeal membrane oxygenation
FDA: Food and Drug Administration
ICU: Intensive care unit
L: Liters
NT-PROBNP: N-terminal-pro B-type natriuretic peptide
O2: Oxygen
SPO2: Oxygen saturation
RECOVERY: Randomized evaluation of COVID-19 therapy SARS-CoV-2 severe acute respiratory syndrome coronavirus 2 WHO World Health Organization
YNHHS: Yale-New Haven Health System

## Acknowledgments

Co-first authors: NB, MVG. Conception and design of work: NB, MVG, SLB, MMT, JEH, CAT, MSD; Acquisition of data: NB, MVG, SLB, AM, MLR, MSD; statistical analysis: FL, YD; interpretation: NB, MVG, SLB, FL, MMT, YD, MSD; Drafting or critically revising for important intellectual content: NB, MVG, FL, SLB, MMT, AM, MLR, YD, JEH, CAT, MSD; Final approval of the version published: NB, MVG, FL, SLB, MMT, AM, MLR, YD, JEH, CAT, MSD; Guarantors ensuring accuracy and integrity of the work: NB, MVG, FL, MSD

The authors thank members of the Yale New Haven Health System COVID-19 Convalescent Plasma Group: Dr. Gregory Buller, Chief of Medicine, Department of Medicine, Bridgeport Hospital; Dr. Herbert Archer, Chief of Medicine and Dr. Gavin McLeod, Infectious Diseases, Greenwich Hospital; Dr. Prakash Kandel, Hospital Medicine, Lawrence and Memorial Hospital; Dr. Mudassar Khan, Hospital Medicine, Westerly Hospital; Dr. Joseph Ladines-Lim, Yale School of Medicine; Dr. Muyi Li and Dr. Alan Zakko, Department of Medicine, Yale School of Medicine and Dr. Kent Owusu, PharmD, Clinical Redesign Consultant Yale New Haven Health for their input in developing criteria for enrolment into the EAP and in enrolling patients into the program. The authors also thank Dr. Peter Peduzzi, Dr. James Dziura, and Dr. Denise Esserman of the Yale Center for Analytical Sciences for their guidance with the analyses.

## Disclaimer

The contents of this publication are the sole responsibility of the authors. The views and opinions expressed in this publication are those of the authors and do not reflect the official policy or position of the US Department of Health and Human Services and its agencies including the Biomedical Research and Development Authority, the Food and Drug Administration, and the National Institutes of Health, as well as any agency of the U.S. government. Assumptions made within and interpretations from the analysis are not reflective of the position of any U.S. government entity.

## Supporting information

**S1 Figure:** Heatmap of change in WHO score for COVID-19 disease severity from time of index to discharge.

**S2 Figure:** Cloud plots showing the distribution of matched propensity scores by patients receiving CCP and not receiving CCP.

A: Matching for early CCP sub-cohort.

B: Matching for late CCP sub-cohort.

**S3 Figure:** Standardized differences for key parameters between CCP recipients and matched unexposed patients.

A: Matching for early CCP sub-cohort.

B: Matching for late CCP sub-cohort.

**S1 Table:** WHO Ordinal Scale for COVID-19 disease severity.

**S2 Table:** Results of propensity score model using logistic regression.

## Notes

All authors declare no conflicts of interest.

### Competing Interest Statement

The authors have declared no competing interest.

### Clinical Trial

This was a medical record review; an observational study. It was not a clinical trial. As such it was not registered with clinicaltrials.gov

### Funding Statement

This project has been funded in whole or in part with Federal funds from the US Department of Health and Human Services; Office of the Assistant Secretary for Preparedness and Response; Biomedical Advanced Research and Development Authority under Contract No. 75A50120C00096. This publication was also made possible by the Yale University Diversity Initiative Faculty Excellence award (to MSD); the CTSA Grant Number UL1 TR000142 (to FL; YD) from the National Center for Advancing Translational Science (NCATS) and the National Institutes of Health; the Immunohematology/Transfusion Medicine Research Training Grant Number T32 HL007974 (to MLR) from the National Heart Lung and Blood Institute (NHLBI); the DeLuca Center for Innovation in Hematology Research at Yale Cancer Center and The Frederick A. Deluca Foundation (to SLB); the Bernard Forget Scholarship in the Section of Hematology and Yale Cancer Center (to SLB).

### Author Declarations

This study was approved by the Yale University Institutional Review Board (HIC#: 2000027871) with an approved waiver of informed consent.

