## Supplemental Material for "Early but not late convalescent plasma is associated with better survival in moderate-to-severe COVID-19"

**Supporting Information**

**Contents**

**S1 Figure:** Heatmap of change in WHO score for COVID-19 disease severity from time of index to discharge.

**S2 Figure:** Cloud plots showing the distribution of matched propensity scores by patients receiving CCP and not receiving CCP.

A: Matching for early CCP sub-cohort.

B: Matching for late CCP sub-cohort.

**S3 Figure:** Standardized differences for key parameters between CCP recipients and matched unexposed patients.

A: Matching for early CCP sub-cohort.

B: Matching for late CCP sub-cohort.

**S1 Table:** WHO Ordinal Scale for COVID-19 disease severity.

**S2 Table:** Results of propensity score model using logistic regression

**S1 Figure:** Heatmap of change in WHO score for COVID-19 disease severity from time of index to discharge.

**Early cohort**

**A	B**

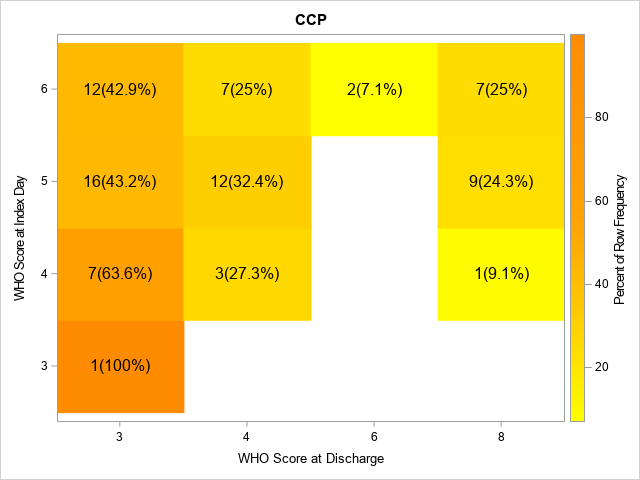

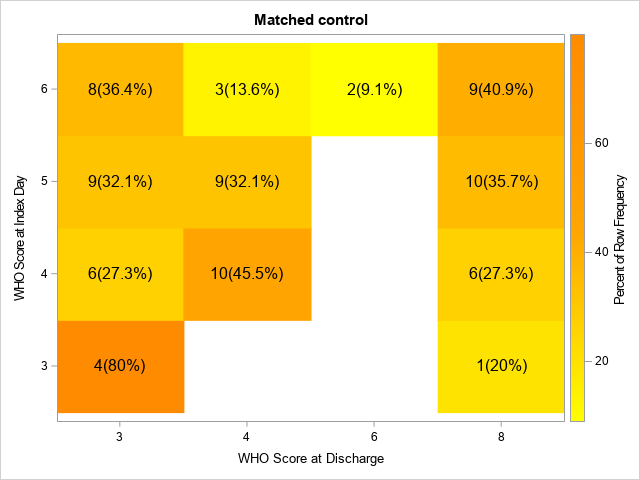

**Late cohort**

**C	D**

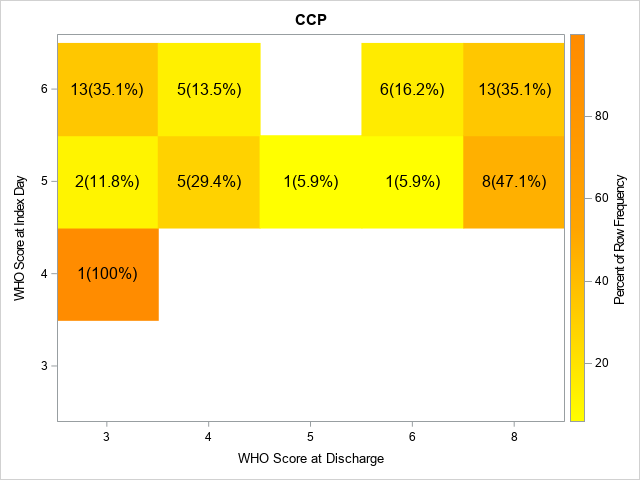

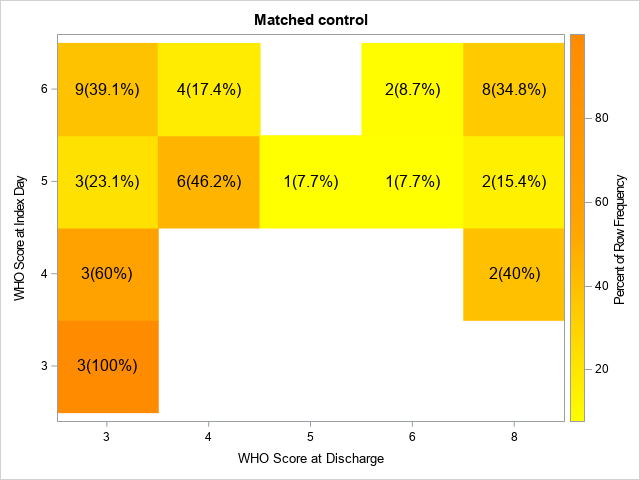

WHO scores of COVID-19 disease severity at discharge (X axis) in patients with the same WHO score at index time (Y axis). Early CCP (A) and late CCP (C) recipients were compared to matched unexposed patients (B, D). In parentheses are the percentages of patients at discharge who shared the same WHO score at index date in each group. Patients in early CCP group were more likely to achieve lower score at the end. For example, 42.9% (12/28) of those who started with score of 6 reached an improved score of 3 as compared to 36.4% in the unexposed group (8/22), and 25% (7/28) of CCP patients starting with 6 ended up with 4 as compared to 13.5% in the unexposed group (3/22), indicating greater odds of clinical improvement in the ordinal scale WHO score at the end (odds ratio for improvement: 1.9, 95% CI 1.1 to 3.3, *P*=0.02, adjusted for WHO score on index day). In contrast, we did not observe same trend in the late cohort, in which the odds ratio for improvement was 0.6 (95% CI: 0.3, 1.3, *P*=0.22).

**S2 Figure**: Cloud plots showing the distribution of matched propensity scores by patients receiving CCP and not receiving CCP.

A: Matching for early CCP sub-cohort

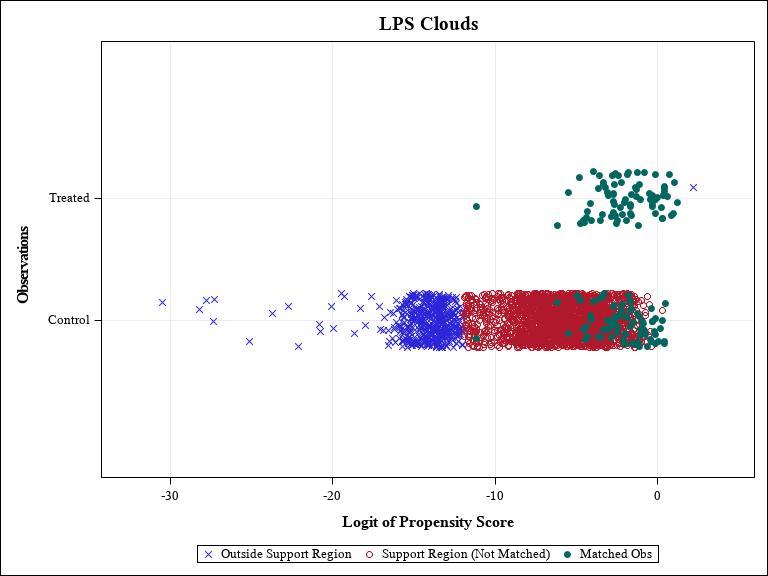

B: Matching for late CCP sub-cohort

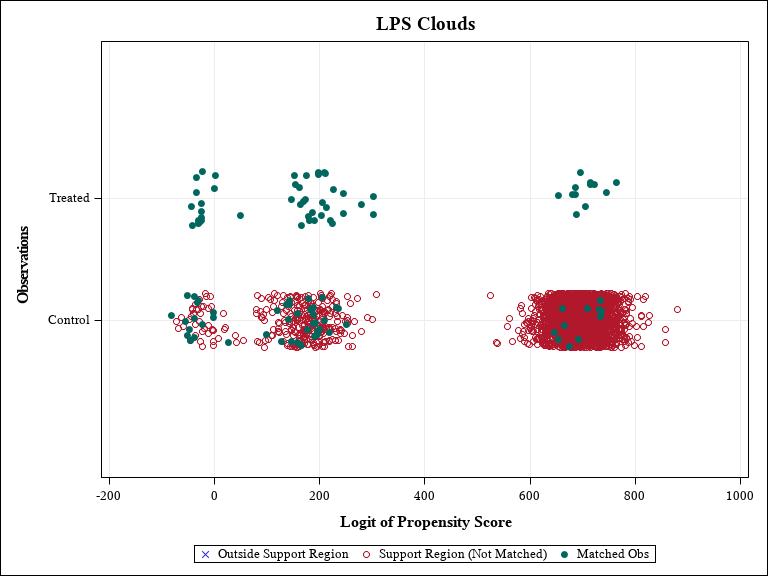

Selection of matched unexposed patients for early CCP (A) and late CCP (B) recipients, respectively compared to unexposed COVID-19 patients that did not receive CCP (n=3217). Blue “X” represents unexposed patients that were outside of the support region for matching based on logit of propensity score. Unexposed patients within the region for selection of matching (red empty circle) were then selected one to one based on similarity of score and represented as green solid circles. Abbreviations: Obs = observations.

**S3 Figure**: Standardized differences for key parameters between CCP recipients and matched unexposed patients.

A: Matching for early CCP sub-cohort

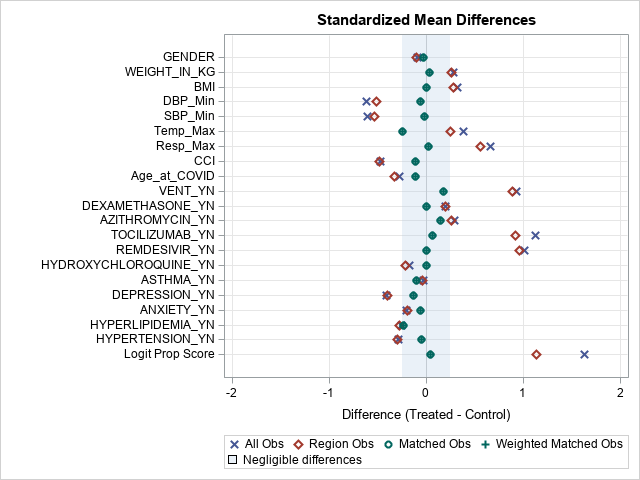

B: Matching for late CCP sub-cohort

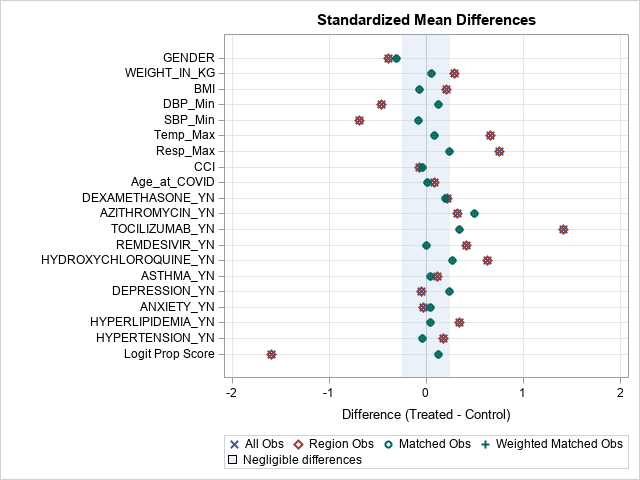

Key parameters were standardized between CCP recipients (green circle) and matched unexposed patients (green plus), compared to all COVID-19 patients outside of the propensity support region (blue X) and within (red diamond). Abbreviations: Obs = observations.

**S1 Table:** WHO Ordinal Scale for COVID-19 disease severity

| **Score** | **Descriptor** | **Patient State** |
| --- | --- | --- |
| 0 | No clinical or virological evidence of infection | Uninfected |
| 1 | No limitation of activities | Ambulatory |
| 2 | Limitation of activities |  |
| 3 | Hospitalized, no oxygen therapy | Hospitalized Mild Disease |
| 4 | Oxygen by mask or nasal prongs |  |
| 5 | Non-invasive ventilation or high-flow oxygen | Hospitalized Severe Disease |
| 6 | Intubation and mechanical ventilation |  |
| 7 | Ventilation and additional organ support: pressors, CRRT, ECMO |  |
| 8 | Death | Dead |

Abbreviations: WHO = World Health Organization

**S2 Table:** Results of propensity score model using logistic regression.

**2a. Propensity score model for early cohort**

| **Predictors for receiving CCP treatment** | **Odds Ratios** | **95% confidence intervals** | | ***P*-value** |
| --- | --- | --- | --- | --- |
| **Smoke** |  |  | | 0.10 |
| Current smoker | 3.09 | 1.17 | 8.12 |  |
| Former smoker | 0.92 | 0.46 | 1.83 |  |
| Do not know | 1.16 | 0.54 | 2.52 |  |
| Never smoke | 1.00 |  |  |  |
| **Worst WHO score within 6 days** |  |  |  | 0.0006^***^ |
| 3 (Hospitalized, no oxygen therapy) | 0.10 | 0.01 | 1.18 |  |
| 4 (Oxygen by mask or nasal prongs) | 0.14 | 0.03 | 0.68 |  |
| 5 (Non-invasive ventilation or high-flow oxygen) | 1.41 | 0.49 | 4.01 |  |
| 6 (Intubation and mechanical ventilation) | 1.00 |  |  |  |
| **Hydroxychloroquine** | 0.46 | 0.21 | 0.98 | 0.04^*^ |
| **Remdesivir within 6 days** | 4.36 | 2.05 | 9.27 | 0.0001^***^ |
| **Remdesivir after 6 days** | 5.52 | 1.90 | 16.07 | 0.002^**^ |
| **Tociluzimab** | 3.62 | 1.49 | 8.85 | 0.005^**^ |
| **Hypertension** | 0.62 | 0.32 | 1.20 | 0.15 |
| **CCI** | 0.89 | 0.78 | 1.02 | 0.11 |
| **Lowest SpO2 within 6 days** | 0.98 | 0.97 | 1.00 | 0.05 |
| **Lowest SBP within 6 days, mmHg** | 1.02 | 0.99 | 1.04 | 0.19 |
| **Lowest DBP within 6 days, mmHg** | 0.97 | 0.94 | 1.00 | 0.08 |
| **Highest Procalcitonin within 6 days, ng/mL** | 0.96 | 0.91 | 1.02 | 0.18 |

**2b. Propensity score model for late cohort**

| **Risk factors** | **Odds Ratios** | **95% confidence intervals** | | ***P*-value** |
| --- | --- | --- | --- | --- |
| **Race** |  |  | | 0.02* |
| White | 1.00 |  |  |  |
| Black or AA | 0.33 | 0.12 | 0.97 |  |
| Other | 1.67 | 0.56 | 4.92 |  |
| **Hispanic** | 0.48 | 0.17 | 1.32 | 0.15 |
| **Age, year** | 1.03 | 0.998 | 1.07 | 0.07 |
| **Weight, kg** | 1.04 | 0.997 | 1.08 | 0.07 |
| **CCI** | 0.88 | 0.76 | 1.02 | 0.08 |
| **ICU** | 4.30 | 1.22 | 15.2 | 0.02^*^ |
| **Ventilation** | 8.71 | 3.08 | 24.7 | <0.0001^***^ |
| **Hyperlipidemia** | 2.22 | 0.95 | 5.19 | 0.07^*^ |
| **Lowest SpO2 within 6 days** | 0.98 | 0.96 | 1.01 | 0.13 |
| **Hydroxychloroquine** | 4.06 | 0.76 | 21.7 | 0.10 |
| **Remdesivir within 6 days** | 2.16 | 0.80 | 5.88 | 0.13 |
| **Tociluzimab** | 8.47 | 1.72 | 41.68 | 0.009^**^ |

Results of propensity score model using logistic regression. Only predictors with *P*< 0.2 were presented. Abbreviations: AA = African American; CCI = Charlson Comorbidity Index; ICU = intensive care unit; kg = kilogram; SpO2 = oxygen saturation; WHO = World Health Organization. *, **, and *** correspond to significant *P*-values for comparison between groups of <0.05, <0.01, and <0.001 respectively.
